# Control Strategies against COVID-19 in China: Significance of Effective Testing in the Long Run

**DOI:** 10.1101/2020.08.22.20179697

**Authors:** Yatang Lin, Fangyuan Peng

## Abstract

The coronavirus disease 2019 (COVID-19) outbreak is reasonably contained in China. In this paper, we evaluated the effectiveness of different containment strategies in halting the pandemic spread in both short- and long-term. We combined a networked metapopulation SEIR model featuring undocumented infections, actual mobility data and Bayesian inference to simulate the counterfactual outbreak scenarios removing each one or a combination of the following three policies in place: i) city lockdowns, ii) intercity travel bans, and iii) testing, detection, and quarantine. Our estimates revealed that 11.4% [95% credible interval (CI): 9.7–13.0%] of the infected cases were unidentified before January 23, 2020. The rate grew to 92.5% [95% credible interval (CI): 85.9–94.5%] in early March, thanks to the boost in coronavirus testing capacity. We show that increasing the detection rate of infections from 11.4% to 92.5% alone would explain 75% of the reduction in infections from a no-policy baseline by March 15, 2020.

The most pronounced policy implication is that city lockdowns appeared to be the more effective intervention in the short-run but effective testing is essential in containing the COVID-19 spread in the long run. By March 15, restoring within-city personal contact to its 2019 level would lead to a 678% growth in infections with all the other interventions remaining unaffected. Removing intercity travel restrictions and effective detection measures would lead to 3% and 477% growth, respectively. Extending the time horizon to July 15, the counterfactual increase in infections would become 581%, 3% and 3.0 *∗* 10^5^% had the three classes of interventions been lifted individually.

**Significance:** The COVID-19 pandemic has become a long-term crisis that calls for long-term solutions. We combined an augmented SEIR simulation model with real-time human mobility data to decompose the effects of lockdown, travel bans and effective testing measures in the curtailment of COVID-19 spread in China over different time horizons. Our analysis reveals that the significant growth in the detection rate of infectious cases, thanks to the expansion in testing efficiency, were as effective as city lockdowns in explaining the reduction in new infections up to mid-March. However, as we extended the analysis to July, increasing the detection rate to at least 50% is the only reliable way to bring the spread under control.

## Introduction

Up to June 30, 2020, more than 10.1 million cases of coronavirus disease 2019 (COVID-19) had been reported, with more than half a million deaths. As the World Health Organization (WHO) declared COVID-19 a pandemic on March 11 2020, the world started preparing to live with COVID-19 as the new normal. All-encompassing lockdowns and travel bans would wreck the global economy in the long run. As a result, countries are looking for middle-ground solutions that would neither dry out national medical resources nor paralyze the economy.

As the first country to sustain a major COVID-19 impact in late December 2019, China has taken unprecedented measures to contain the spread of disease, including physical distancing, travel bans, testing, case identification, and quarantine. The measures have appeared to be effective: after March 15, 2020, without considering the imported cases from other countries, less than 100 new cases were reported per day in China. On April 8, China lifted its 76-day lockdown of Wuhan. The country was reopening businesses and schools gradually.

So what is there to be learned from China’s COVID-19 experiences? Our study aimed to quantitatively evaluate the contribution of the three major types of policies implemented for successful containment of COVID-19: i) city lockdown aiming to reduce within-city contact, ii) cross-city travel restrictions, and iii) effective ways to test and isolate infected persons. Importantly, we followed the full trajectory of COVID-19 in China from January 10, two weeks before the drastic lockdown of Wuhan on January 23, to March 15, when the pandemic was contained. The long period study period enabled us to *estimate* rather than simulate the policy effects.

We based our analysis on a Susceptible-Exposed-Infected-Recovered (SEIR) model adapted from (*1*). We explicitly modelled the testing, detection and quarantine process by categorizing infections into documented and undocumented ones and imposing different parameters of transmission between the two categories. A radical ramping up of testing could turn a majority of new infections into the less transmissive class and crucially slow down the spread. We estimated the model for five sub-periods from January 10 to March 15, 2020, and mapped the changes in key parameters to observed changes in different non-pharmaceutical interventions in reality.

The impacts of city lockdown and intercity travel bans were modelled as reductions in within- and intercity mobility, which affects the probability of contact and disease transmission across individuals. We obtained daily real-time mobility data from Baidu Migration, a travel map offered by the largest Chinese search engine. To derive a counterfactual scenario where the restrictions on with- and intercity mobility had never been implemented, we aligned the 2019 and 2020 Baidu Migration mobility data on the basis of relative timing to the Spring Festival. For example, we assumed that without intercity travel bans, the counterfactual number of travellers between city pairs on January 23, 2020 (2 days before Spring Festival) would be the same as the observed number of travellers on February 3, 2019. Similarly, reduction in intercity mobility from the 2019 baseline level was used to estimate the effects of city lockdown on contact reductions.

A first glance at the mobility data in 5 and 4 revealed significant reductions in population inflow into Hubei cities after January 22, when evidence on human-to-human transmission was revealed. The trend never recovered up to mid-March. The flow into non-Hubei cities also experienced a sudden drop around the same time, but gradually increased after the end of February, when the first wave of pandemic was under control. Similarly, the within-city mobility plunged after January 22 and stayed at a very low level for Wuhan until the end of March. The within-city mobility for cities in other provinces such as Chongqing also decreased significantly around the same period, but gradually caught up to its 2019 level by the end of March, when lockdown regulations had been lifted.

## Results and Discussion

Our estimates revealed that the detection rate of infections grew from 11.3% [95% credible interval (CI): 9.7–13.0%] before January 23, to 92.5% [95% credible interval (CI): 85.9–94.5%] in early March, accompanied by a significant reduction in the transmission rate and the length of infectious period of confirmed patients, evidence consistent with substantial improvement in testing and treatment capabilities.

A direct comparison across the three groups of control methods is presented in Table 2 and Figure 6. We found that drastic suppression measures, such as city lockdowns, were most effective in the short run. In the counterfactual scenario had we lifted city lockdowns after January 23, the cumulative number of infections by February 29 would have been 648% of the reality. Comparatively, keeping the detection rate and transmission parameters at the baseline (before January 23) level produced an additional 69% infections. Restoring intercity travel flows to the 2019 level would lead to a threefold growth in infected cases out of Wuhan but would have limited effects on Wuhan. The three containment measures had strong complementary effects: lifting all three interventions, the number of cases would have been 65-fold higher by February 29, quite close to the estimates presented by (*2*).

**Table 1:** Best-fit model estimates for key epidemiological parameters

| Paramter | 10 January–23 January | 24 Jan–2 Feb | 3 Feb– 12 Feb | 13 Feb–22 Feb | 23 Feb–15 Mar |
| --- | --- | --- | --- | --- | --- |
| Transmission Rate $\beta^T$ (Reported) | 0.05 (0.03,0.08) | 0.02 (0.01,0.03) | 0.02 (0.02,0.05) | 0.02 (0.01,0.03) | 0.01 (0.01,0.02) |
| Transmission Rate $\beta^{NT}$ (Unreported) | 0.25 (0.24,0.25) | 0.24 (0.23,0.24) | 0.24 (0.23,0.28) | 0.24 (0.23,0.26) | 0.24 (0.23,0.24) |
| Mobility Probability | 0.1 (0.09,0.12) | 0.92 (0.9,0.94) | 0.73 (0.41,0.91) | 0.69 (0.25,0.9) | 0.49 (0.06,0.92) |
| Latent Period | 4.57 (4.48,4.65) | 4.67 (4.57,4.74) | 4.41 (2.46,4.7) | 3.7 (2.32,4.66) | 3.79 (3.03,4.41) |
| Reported Rate | 0.11 (0.1,0.13) | 0.24 (0.13,0.68) | 0.86 (0.24,0.94) | 0.92 (0.12,0.95) | 0.94 (0.86,0.95) |
| Infection Period (Reported) | 1.82 (1.43,2.24) | 1.42 (1.14,2.3) | 1.53 (1.13,2.44) | 1.31 (1.1,2.05) | 1.09 (1.06,1.55) |
| Infection Period (Unreported) | 3.4 (3.27,3.68) | 3.32 (3.2,3.63) | 3.44 (3.22,4.03) | 3.32 (3.2,3.6) | 3.3 (3.22,3.76) |
| Relative reported rate (Wuhan) | 0.01 (0.01,0.01) | 0.01 (0,0.04) | 0.07 (0.01,0.09) | 0.53 (0.05,0.68) | 0.83 (0.56,0.87) |

Notes: This table shows best-fit model posterior estimates of the median and 95% confidence intervals for key epidemiological parameters derived from 200 simulation of the model.

**Table 2:** Alternative Scenarios: Lifting each Control Method

|  | No Control Method<br>(a) | No Travel Ban<br>(b) | No Lockdown<br>(c) | No Testing Treatment<br>(d) |
| --- | --- | --- | --- | --- |
| **Panel A: National** |  |  |  |  |
| *Until 7.15* |  |  |  |  |
| No. of cases | $1.22 \times 10^9$ | $3.84 \times 10^5$ | $2.54 \times 10^6$ | $1.14 \times 10^9$ |
| simulated cases/actual case | 3274.93 | 1.03 | 6.81 | 3069.81 |
| *Until 3.15* |  |  |  |  |
| No. of cases | $1.72 \times 10^8$ | $3.84 \times 10^5$ | $2.52 \times 10^6$ | $2.15 \times 10^6$ |
| simulated cases/actual case | 462.62 | 1.03 | 6.78 | 5.77 |
| *Until 2.29* |  |  |  |  |
| No. of cases | $2.41 \times 10^7$ | $3.84 \times 10^5$ | $2.41 \times 10^6$ | $6.28 \times 10^5$ |
| simulated cases/actual case | 64.70 | 1.03 | 6.48 | 1.69 |
| **Panel B: Hubei** |  |  |  |  |
| *Until 7.15* |  |  |  |  |
| No. of cases | $5.99 \times 10^7$ | $3.47 \times 10^5$ | $2.50 \times 10^6$ | $2.21 \times 10^7$ |
| simulated cases/actual case | 168.81 | 1.00 | 7.03 | 62.17 |
| *Until 3.15* |  |  |  |  |
| No. of cases | $4.04 \times 10^7$ | $3.47 \times 10^5$ | $2.48 \times 10^6$ | $5.48 \times 10^5$ |
| simulated cases/actual case | 113.89 | 1.00 | 6.99 | 1.54 |
| *Until 2.29* |  |  |  |  |
| No. of cases | $1.40 \times 10^7$ | $3.46 \times 10^5$ | $2.37 \times 10^6$ | $4.66 \times 10^5$ |
| simulated cases/actual case | 39.36 | 1.00 | 6.69 | 1.31 |
| **Panel C: Other Citiy** |  |  |  |  |
| *Until 7.15* |  |  |  |  |
| No. of cases | $1.16 \times 10^9$ | $3.77 \times 10^4$ | $4.13 \times 10^4$ | $1.12 \times 10^9$ |
| simulated cases/actual case | 67139.22 | 2.19 | 2.39 | 64909.19 |
| *Until 3.15* |  |  |  |  |
| No. of cases | $1.32 \times 10^8$ | $3.77 \times 10^4$ | $4.09 \times 10^4$ | $1.60 \times 10^6$ |
| simulated cases/actual case | 7638.59 | 2.18 | 2.37 | 92.81 |
| *Until 2.29* |  |  |  |  |
| No. of cases | $1.01 \times 10^7$ | $3.74 \times 10^4$ | $3.88 \times 10^4$ | $1.62 \times 10^5$ |
| simulated cases/actual case | 589.17 | 2.18 | 2.26 | 9.46 |

*Notes:* This table shows alternative counterfactual scenarios lifting each one of the three control measures (lockdown, intercity travel ban and improved testing efficiency) and all three of them together (no control measure). Column (a) shows scenario without any control measure in place since January 23. Column (b),(c) and (d) present counterfactual number of infectious cases under scenarios that lift intercity travel restriction, lockdown and testing and treatment measures, respectively. Results in this table are the mean value derived from 200 simulations.

**Table 3:** Alternative Scenario: different testing efficiency

| Detection Rate | 30% | 50% | 70% | 90% |
| --- | --- | --- | --- | --- |
| **Panel A: Until 3.15** |  |  |  |  |
| *Scenario 1: No lockdown ever* |  |  |  |  |
| No. of cases | $1.55 \times 10^7$ | $4.28 \times 10^6$ | $2.58 \times 10^6$ | $2.05 \times 10^6$ |
| simulated cases/actual cases | 41.5 | 11.5 | 6.9 | 5.5 |
| *Scenario 2: Lockdown since Jan 23* |  |  |  |  |
| No. of cases | $6.28 \times 10^5$ | $4.70 \times 10^5$ | $4.40 \times 10^5$ | $4.27 \times 10^5$ |
| simulated cases/actual cases | 1.74 | 1.30 | 1.22 | 1.18 |
| **Panel B: Until 7.15** |  |  |  |  |
| *Scenario 1: No lockdown ever* |  |  |  |  |
| No. of cases | $6.95 \times 10^8$ | $7.72 \times 10^6$ | $2.68 \times 10^6$ | $2.07 \times 10^6$ |
| simulated cases/actual cases | 1866.9 | 19.4 | 7.2 | 5.6 |
| *Scenario 2: Lockdown since Jan 23* |  |  |  |  |
| Cases | $2.22 \times 10^8$ | $5.01 \times 10^5$ | $4.51 \times 10^5$ | $4.41 \times 10^5$ |
| simulated cases/actual cases | 595.14 | 1.38 | 1.22 | 1.18 |
| *Scenario 3: Lockdown from Jan 23 to Mar 15, no lockdown after Mar 15* |  |  |  |  |
| No. of cases | $2.89 \times 10^8$ | $4.97 \times 10^5$ | $4.40 \times 10^5$ | $4.27 \times 10^5$ |
| simulated cases/actual cases | 799.59 | 1.38 | 1.22 | 1.18 |

*Notes:* This table presents alternative counterfactual scenarios with different detection rate after March 15. We let the detection rate to increase from the initial level before January 23, gradually to the designated level from January 23 to March 15. We simulate different lockdown scenarios and set the inner city mobility to follow the level on the same lunar calendar date in 2019 in the counterfactual case of no lockdown. The three scenarios are (1) No lockdown ever from January 10 to July 15; (2) Lockdown from January 10 to July 15; (3) Lockdown from January 10 to March 15, no lockdown from March 15 to July 15. Results in this table are the mean value derived from 200 simulations. After March 15 when the daily mobility data were no longer available, we use the average migration and inner-city flow intensity from March 9 to March 15 (one week before March 15) as a proxy.

However, as we extended the time horizon of the analysis, the cumulative effects of different control measures reversed. City lockdown by itself was not sufficient to bring the spread under control. In the counterfactual scenario with both city lockdowns and travel bans in place, leaving the detection rate at 30% would predicted 2.22 *×* 10^8^(95% credible interval (CI): 6.6 *×* 10^7-^3.2*×*10^8^) cases by July 15, 595 times the factual number of infections, compared to 1.2 times if the detection rate was set at 70%. On the contrary, with efficient testing, tracing, and treatment, we could afford to relax restrictions on both within-city and intercity mobility. In case the detection rate at 70% could be upheld, restoring within-city and intercity mobility to 2019 level since the beginning of the outbreak would only lead to a 7.2-fold growth in infected cases.

Perhaps the strongest policy implication emerge from our evidence is that the detection rate of infectious cases has to be higher than 50% to bring the pandemic under contril in the presence of strict city lockdown at the beginning of outbreak, and higher than 70% without. (*3*) estimated the detection rate of COVID-19 infections across 10 countries based on a demographic scaling model and age-specific infection fatality rates (IFRs). By mid-May, the estimated detection rate was less than 20% in Italy, 45% for the U.S. and 55% for Germany, which could explain the diverging performance in COVID-19 control across these countries. Consequently, investments in testing capacity and contact-tracing systems should be placed in a high priority to prevent ongoing secondary outbreaks of COVID-19 or similar future outbreaks of other emergent infectious diseases.

## Materials and Methods

### Model setup and estimation

We modelled the transmission of COVID-19 using a metapopulation Susceptible-Exposed-Infected-Recovered (SEIR) framework, which can flexibly generate patterns of spatial transmission (See Figure 1 for an illustration of the model structure). We traced the spatial spread of COVID-19 across cities with mobility data from Baidu Migration, a data service provided by the largest Chinese search engine. Baidu Migration collects information on population mobility from real-time location records of smartphones that use its mapping app. The platform reported bilateral migration indices for 36057 city pairs per day for 365 Chinese cities between January 12 and March 26 in 2019, and between January 1 and March 15 in 2020. It also published daily within-city mobility data for each city during the sample period. The period covers the annual “Chunyun” (Spring Festival travel season) mass migration cycle.

**Figure 1:**
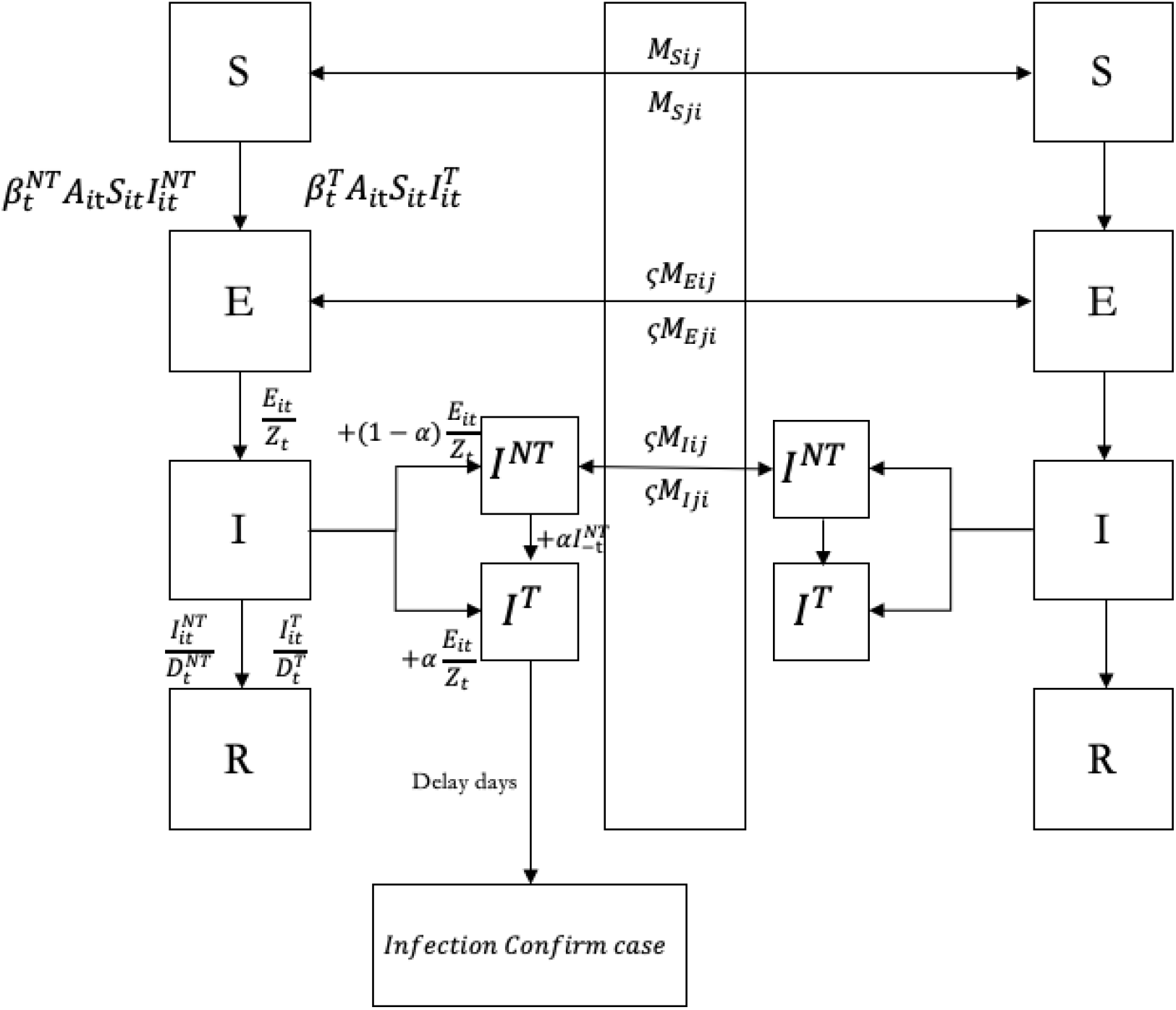
SEIR Model.

*Notes:* Graphical scheme representing the interactions among different stages of infection in SEIR Model. In this model, 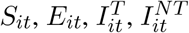 and *N_it_* denote the susceptible, exposed, detected infected, undetected infected and total population in city *i* and time 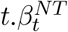 and 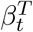 are the rate of transmission for undetected infected individuals and undetected infected individuals, respectively. *α_t_* is the testing rate in time t.*Z_t_* denotes latency period through which patients switch from exposed stage to infection stage.*A_it_* is within-city population flow in city i in time t. Spatial spread of the disease is governed by the daily number of people travelling from city j to city i in time t(*M_ijt_*).

To evaluate the role of testing, detection and post-diagnosis quarantine, we divided infections into documented and undocumented cases (*I^T^* and *I^NT^* in Figure 1). The two types of infections had different rates of transmission (*β*) and infectious period (1*/λ*). According to the model, extensive testing and detection (higher detection rate *α*) would help reduce the transmission risk of infected persons as they would be quickly isolated and treated.

To capture the changes in epidemiological characteristics of the outbreak over time, especially after January 23, when serious control measures were implemented, and after Feb 08, when industries were gradually re-opened, we divided the full sample from January 1 and March 15 into five sub-periods: January 10 to January 23, January 24 to February 2, February 3 to February 12, February 13 to February 22, and February 23 to March 15. We estimated the key parameters (*α, βdocumented, βundocumented, γdocumented, γundocumented*) separately for each sub-period, and mapped the changes to factually improvements in control measures. To better characterize the overwhelmed testing capacity at the early outbreak in Wuhan, we allow the detection rate *α* to differ between Wuhan and the rest cities.

We also gathered data on the time gaps between illness and case report date from (*4*) to estimate a time-to-event delay function. For each of a new case, we drew a reporting delay from a gamma distribution. The purpose is to bridge the synthetic and observed outbreaks and improve the model fit.

### Validation of the model-inference framework

We estimated key parameters of the model using an iterated filter-ensemble adjustment Kalman filter (IF-EAKF) approach (*1*). The framework identified the maximum likelihood estimates of key parameters listed in Table 1. The transmission rate (*β*) for undocumented infections was less than 22% of that for documented cases from January 10 to January 23, 2020. The ratio further dropped to 11% after February 3. The infectious period for positive cases also dropped from 1.827 days in late January to 1.156 days in early March. Both effects could be attributable to improvements in the treatment capacity and practices in managing confirmed patients. A notable example is the introduction of Fangcang shelter hospitals, a rapidly-constructed and low-cost medical infrastructure providing basic isolation, triage, medical care, monitoring and referral services to clinically confirmed patients. The development of Fangcang hospitals initiated on February 5, 2020 significantly reduced intra-family transmission associated with home isolation and was considered a critical move toward balancing the strained medical system in the Hubei province(*5*). Meanwhile, the detection rate *α* proliferated from less than 1% in Wuhan before January 23 to more than 70% in early March, as shown in Figure 3. The detection rate in other cities grew steadily from less than 10% at the onset of the pandemic to more than 90% in early March. The estimated percentage of undocumented cases at the beginning of the outbreak in Wuhan was slightly lower than the ones reported by (*1*) and (*6*), primarily driven by differences in sample period definition. Our estimated basic reproductive number, R0, is 3.88 [95% credible interval (CI): 3.70–4.32], consistent with other recent estimates in similar settings((*7*),(*8*),(*9*),(*10*)).

**Figure 2:**
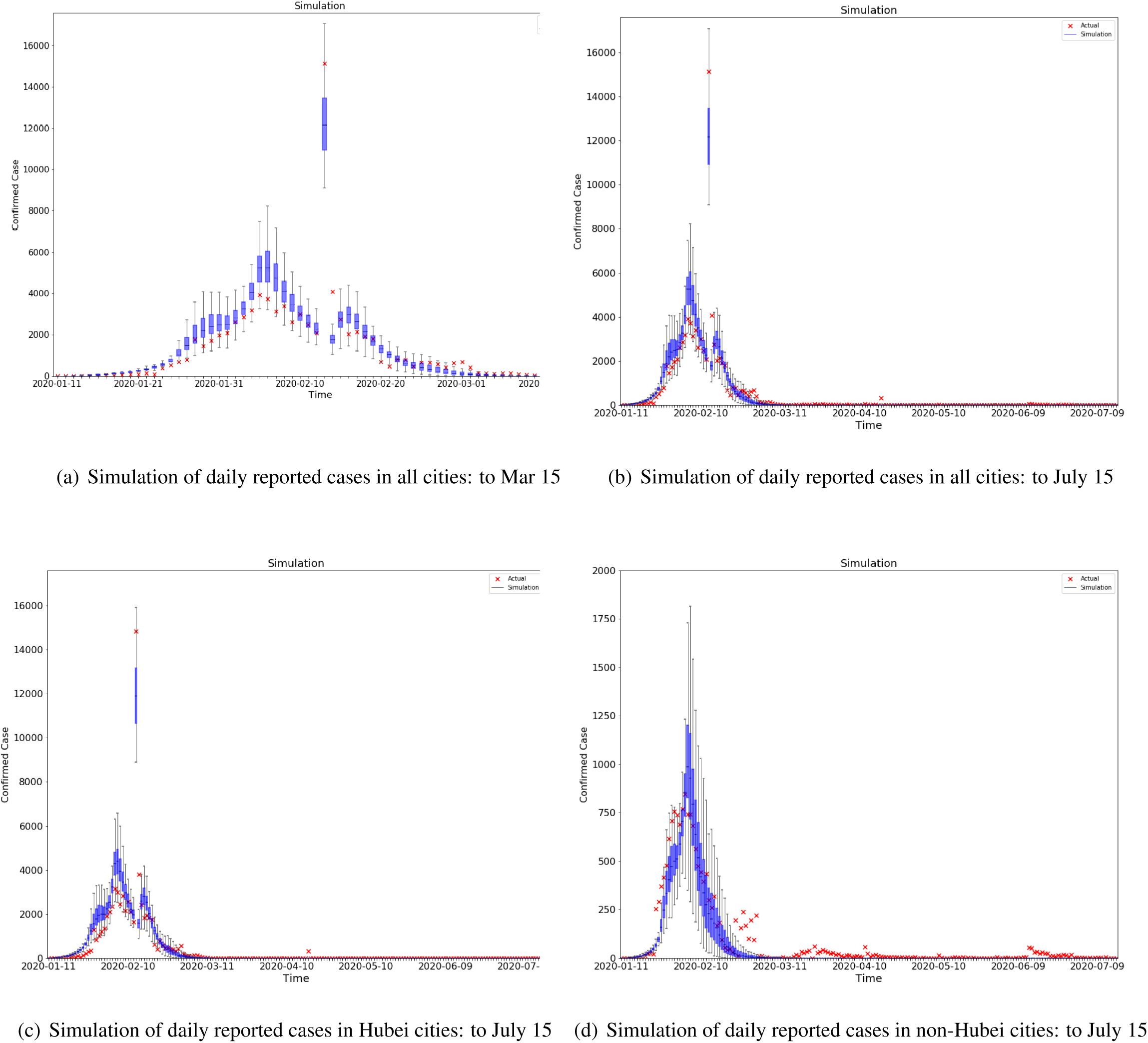
Model fit and out-of-sample prediction.

*Notes:* These graphs compare the daily official reported number of cases to the simulated ones from 200 simulations using the fully-estimated model in all cities from Jan 10,2020 to March 15,2020 (a), from Jan 10,2020 to July 15,2020 (b), in cities from Hubei province from Jan 10,2020 to July 15,2020 (c), in cities outside Hubei province (d) from Jan 10,2020 to July 15,2020. The orange x deontes the number of daily reported cases. The blue box and whiskers show the median, interquartile range(IQR), and 1.5IQR derived from 200 simulations using the fitting model with parameters estimated from 1 *R*^2^ = 0.86-0.97.

**Figure 3:**
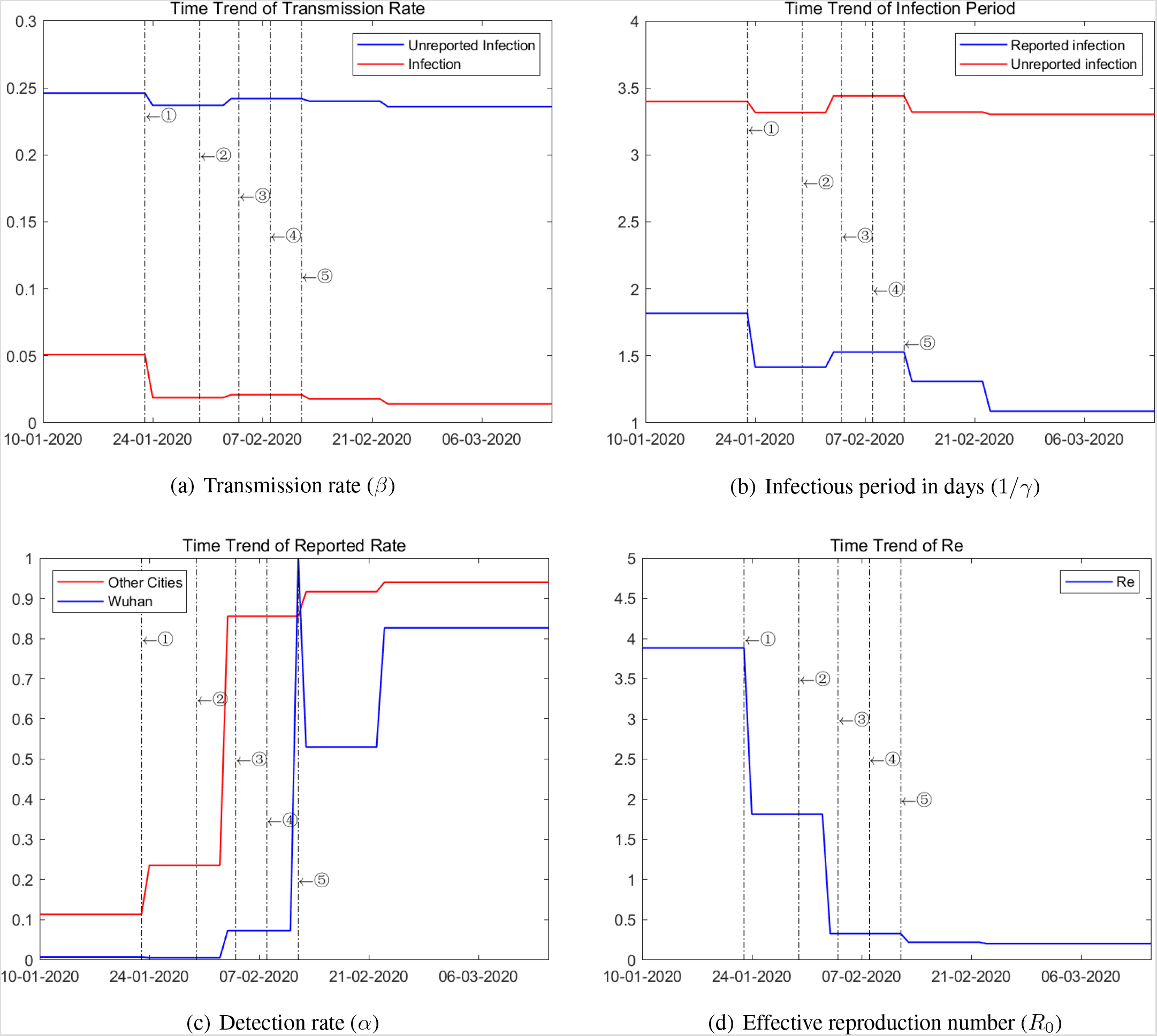
Time Trend of Parameters.

Notes: These graph plot the changes in parameter value estimated over five subperiods: January 10 to January 23, January 24 to February 2, February 3 to February 12, February 13 to February 22, and February 23 to March 15.

**Figure 4:**
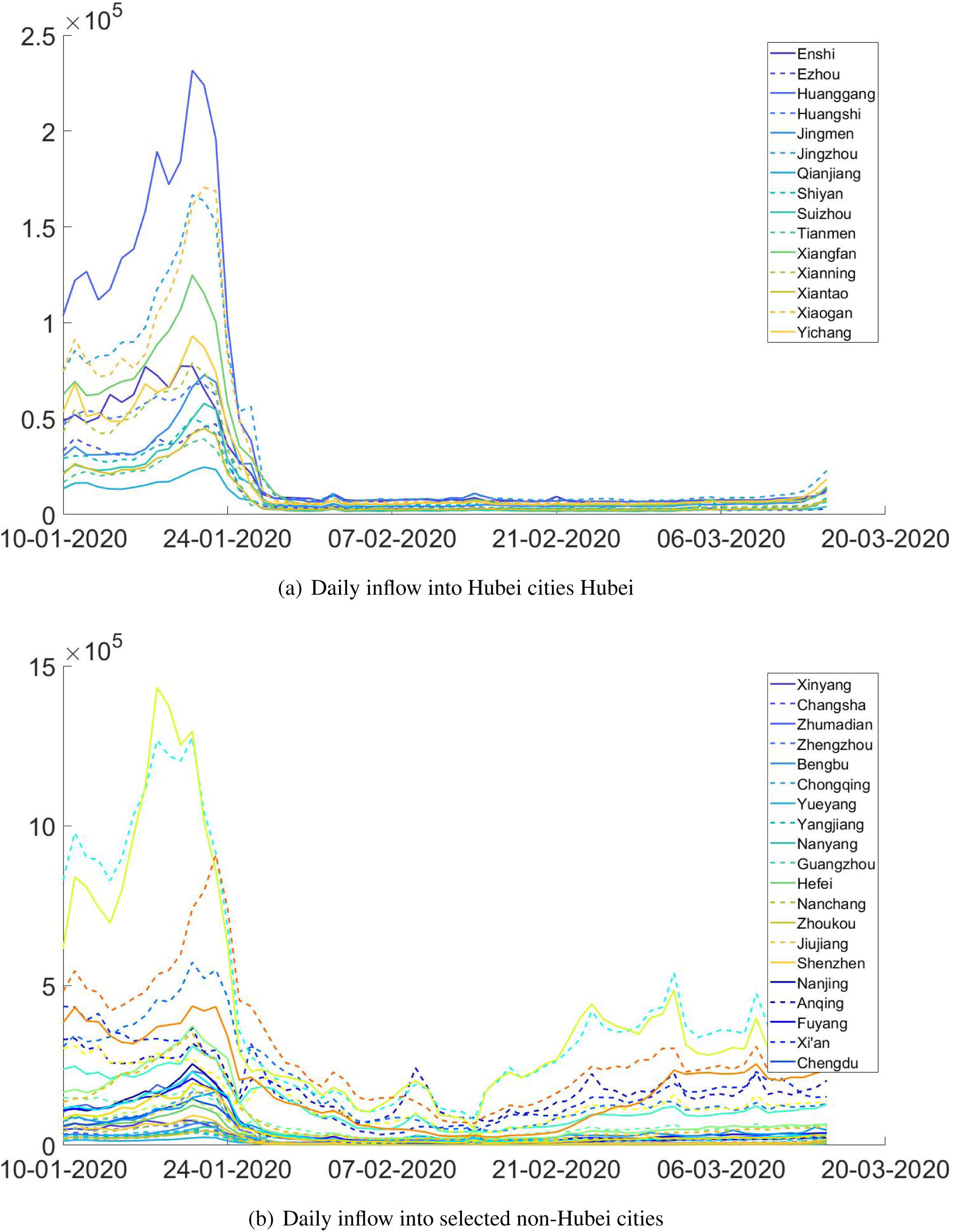
Inter-city Population Flows.

*Notes:* These graphs plot the size of daily aggregate population inflow into cities in Hubei(a) and selected cities out of Hubei (b).

**Figure 5:**
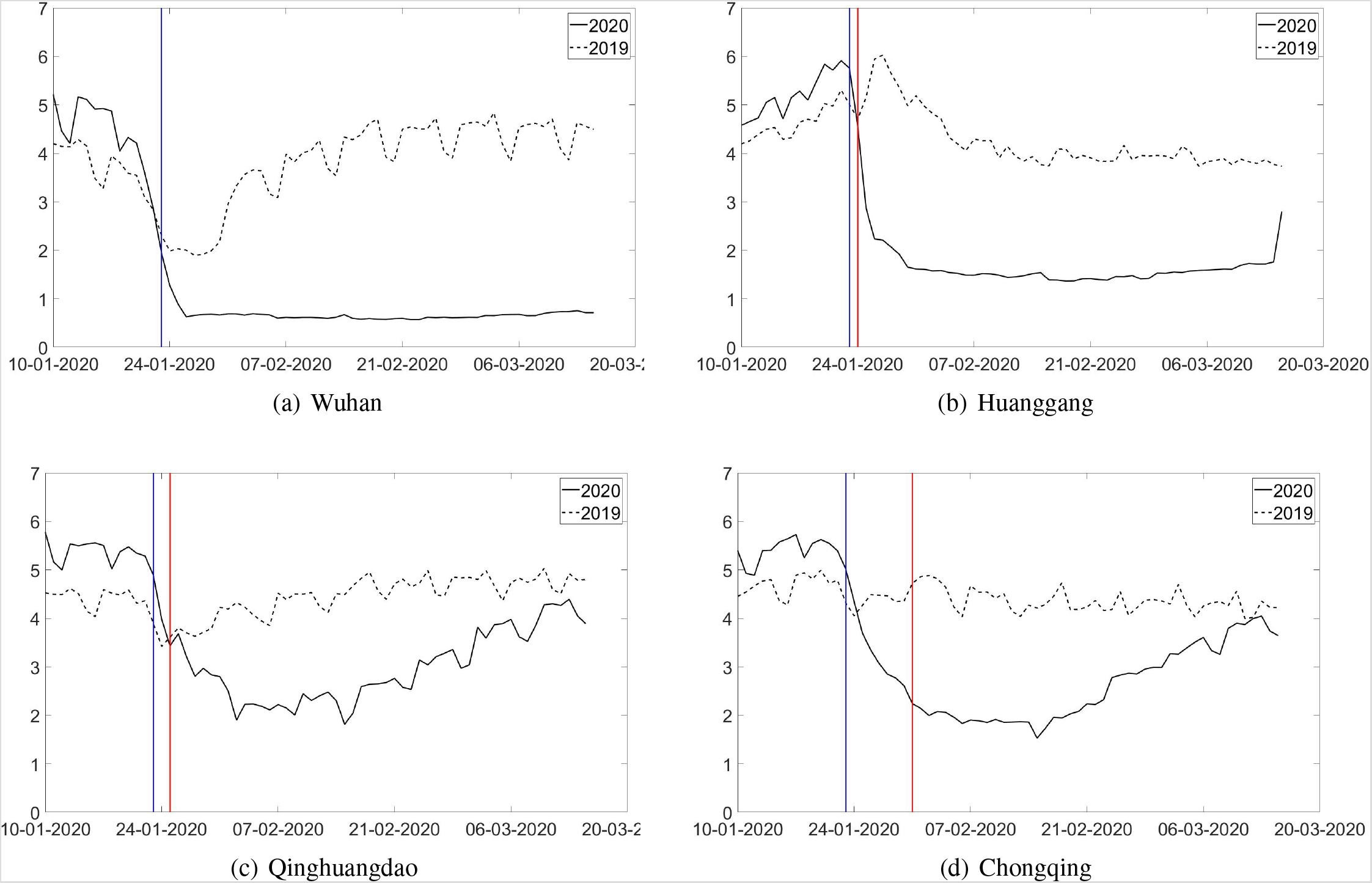
Within-City Mobility.

*Notes:* These graphs plot daily variations in within-city mobility in Wuhan(a), Huanggang(b), Qinghuangdao(c) and Chongqing(d) in 2019 and 2020. Blue Line indicates the date of Wuhan lockdown (Jan 23,2020) and red line indicates the date of each city’s own lockdown.

**Figure 6:**
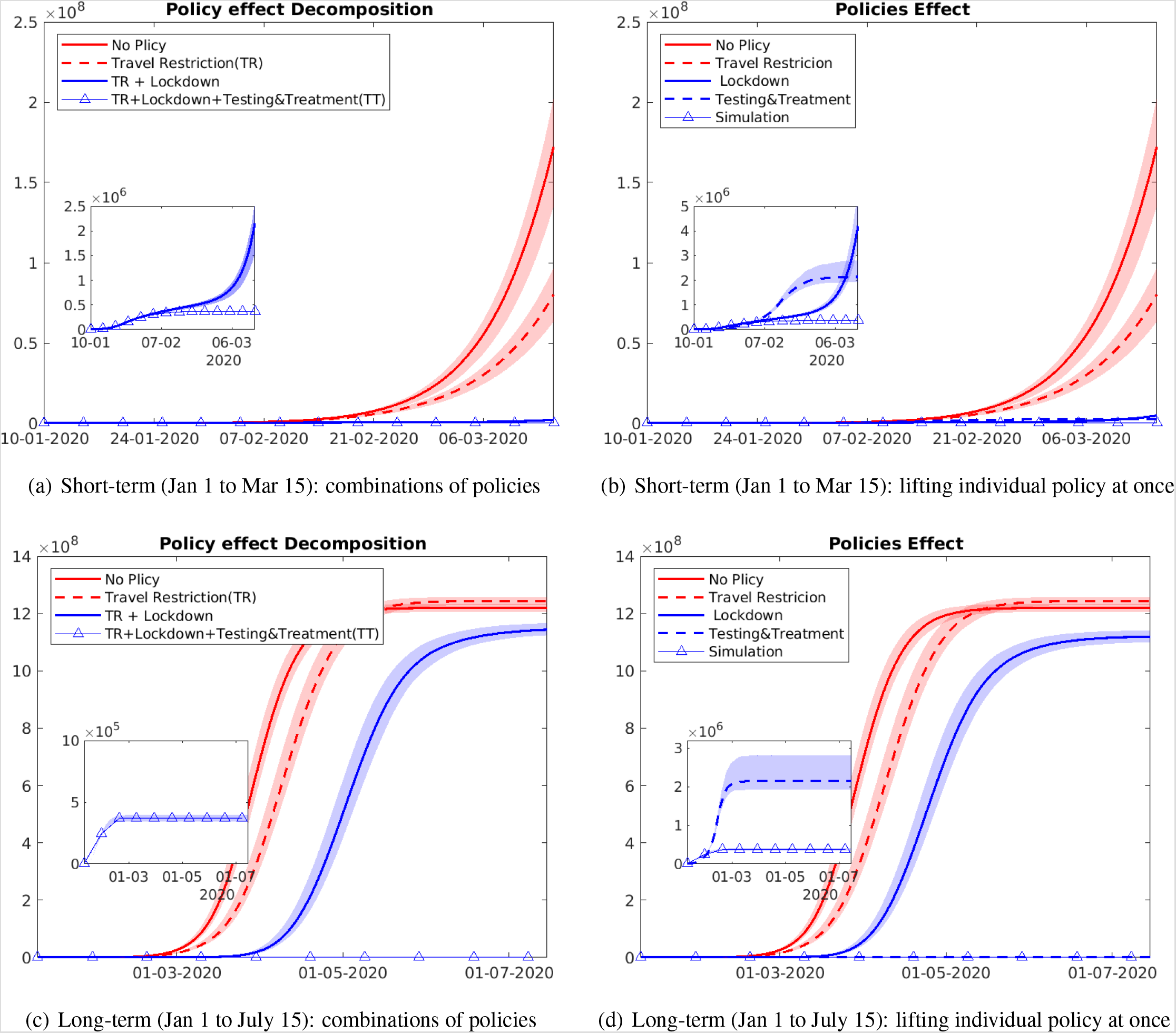
Alternative scenarios: Lifting combinations of different policies.

*Notes:* This figure plots the trends in observed and counterfactual number of cumulative infections under several alternative scenarios from Jan 1,2020 to Mar 15,2020, and from Jan 1 to July 15. Data are presented as the median (solid line) and IQR (shading) of estimates (200 simulations).”No Policy” denotes the scenario in which no anti-contagion policies have been put in place. “Travel Restriction” denotes the case where only intercity travel bans were implemented after January 23.”Lockdown” denotes the scenario in which only city lockdown was implemented after January 23. “Testing & Treatment” denotes the scenario in which the detection rates of infections were fixed at the levels before January 23. “Simulation” denotes the baseline scenario in which all three policies have been implemented from January 23 to March 15. “TR+Lockdown” denotes the scenario in which both intercity travel restrictions and city lockdown were implemented. “TR+Lockdown+Testing&Treatment(TT)” denotes the baseline scenario in which all three policies have been implemented from January 23 to March 15.

**Figure 7:**
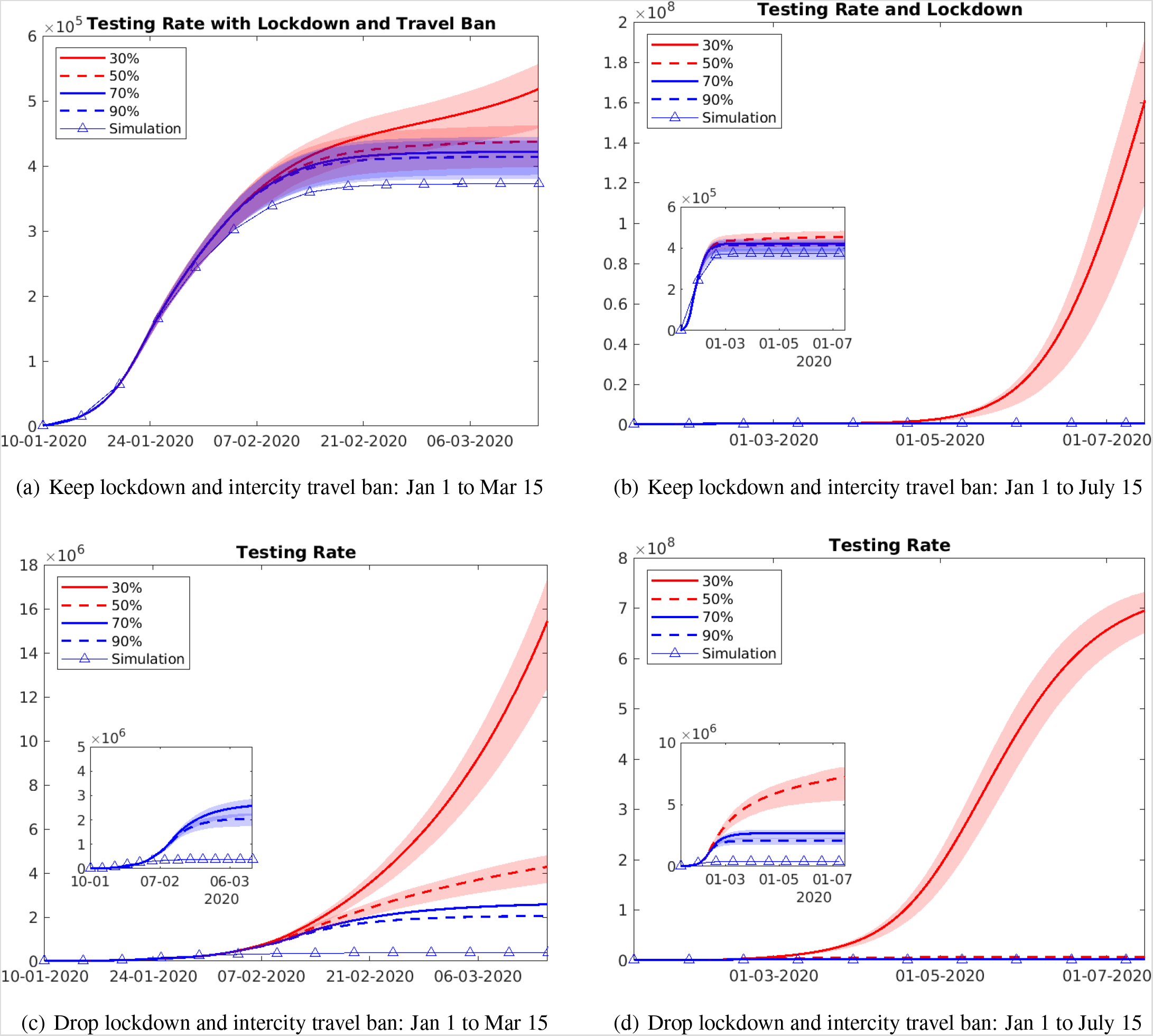
Alternative scenarios with difference testing efficiency.

*Notes:*This figure plots the trends in observed and counterfactual number of cumulative infections under alternative infection detection rates from Jan 1,2020 to March 15,2020 and from Jan 1,2020 to July 15,2020. Data are presented as the median (solid line) and IQR (shading) of estimates (200 simulations). In the scenario presented in the upper panel, we keep the other two policies (lockdown and intercity travel ban) in place, and adjust the detection rate of infections to be 30%, 50%, 70% and 90% respectively. In the lower panel, we drop the lockdown and travel ban policies. The other parameters such transmission rate *β* and infectious period 1*/γ* are the same as in the baseline.

**Figure 8:**
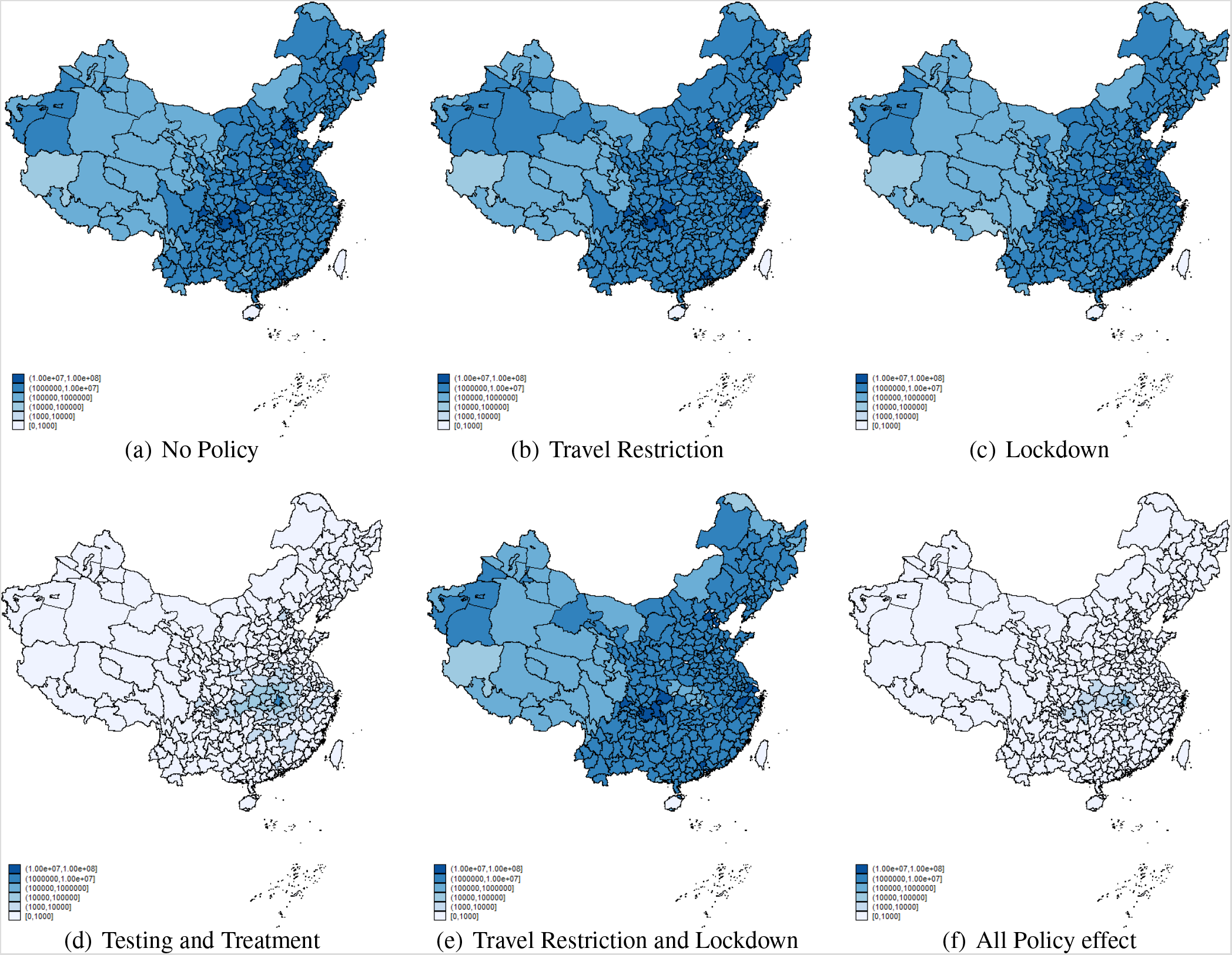
Spatial distribution of infections under alternative scenarios: July 15, 2020.

*Notes:* This graph shows the spatial distribution of cumulative infectious cases on July 15, 2020 under different scenarios.Figure (a) plots number of total reported case distribution without any policy implemented. Figure (b),(c),(d) and (e) shows the cumulative number of cases under travel restriction, lock-down, testing and treatment and the combination of travel restriction and lock-down,respectively. Figure (f) simulates the baseline distribution of infections with all three policies in place.

We present simulations of reported cases generated by the model inFigure 2. The simulation matches well with the observed outbreaks for both the Hubei province and the rest of China, even though we did not target at matching the two subgroups separately. The explicit modelling of reporting activities also makes our model more flexible to account for surges in reported cases as a result of changes in case identification criteria. Estimation of epidemiological models on China in previous articles ((*2*),(*11*),(*12*)) usually stops before February 13. On that day, China revised the case definition in Hubei to include patients who met the clinical criteria in the absence of a positive PCR test, purportedly to clear the backlog of COVID-19 tests. To account for this outlier, we manually set *α* = 1 for Wuhan on February 13, 2020.

As is clear from Figure 2, the surge was well simulated in our model. This operation allowed us to extend our analysis to March 15, the last day when Baidu Migration mobility data were made available. An obvious benefit of extending the period of study is that we could look at changes in key parameters in response to new policy changes after February 13, including the opening of temporary hospitals and tentative re-opening of industries. Assuming that the control measures were kept at similar levels from March 15 to July 15, we borrowed the parameter estimates and mobility measures from the last period of our sample (February 23 to March 15) to perform an out-of-sample model validation exercise. The predicted cumulative infections on July 15 is only 17% higher than the actual ones, a sign that our model could capture the intensity of containment policies well.

## Data Availability

The data will be made available shortly.

## Supplementary Materials

### Model Framework

#### Initialization

The transmission model adopts the following metapopulation structure:

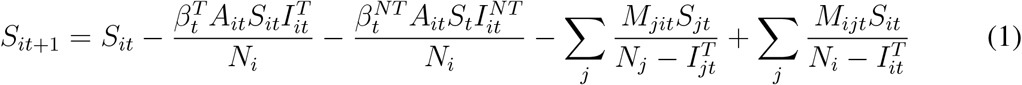

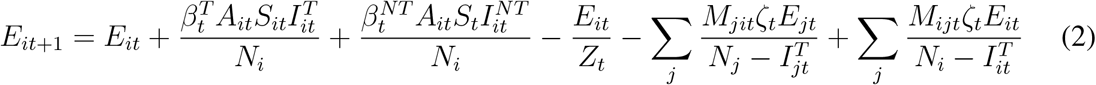

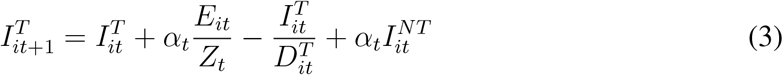

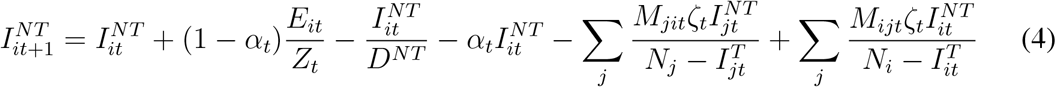

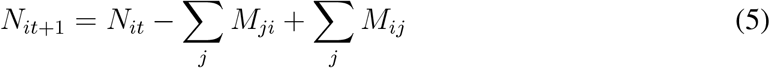

where 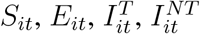 and *N_it_* are the susceptible, exposed, detected infected, undetected infected and total population in city *i* and time *t*. The interactions among different stages of infection are visually represented in Fig. The parameters are defined as follows:

- 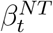 demotes the rate of transmission for undetected infected individuals. *A_it_* denotes within-city population mobility. Their product is going to be the probability of transmission among infected persons. 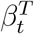 is the rate of transmission for undetected infected individuals. Typically 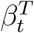 should be smaller than 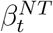, provided that confirmed infected persons are properly quarantined.
- *α_t_* is the testing rate in time t, which is defined as the ratio between the number of documented infections in time t and the sum of cumulated undetected patients carried over from last period and new patients in time t.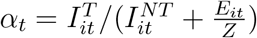
- *Z_t_* denotes latency period through which patients switch from exposed stage to infection stage. *D_t_* is the infectious period that patients could infect the susceptible population. 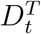 is the infectious duration for detected infections while 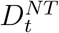 is for undetected infections. *D_t_* is typically lower than 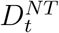 provided that detected infected individuals would be properly treated and become well sooner.
- *A_it_* is within-city population flow in city i in time t. Spatial spread of the disease is governed by the daily number of people travelling from city j to city i in time t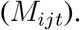 To capture the fact that exposed and undetected individuals might travel less to other cities, likely because of the illness of family members or voluntary avoidance behavior following the reports of epidemic hot spots, we add a multiplicative factor, *ζ* smaller than one. We further assume that individuals in the tested group who have been admitted by local hospital do not move between cities.

In this model, the effective reproduction number (*R*_0_) is calculated as 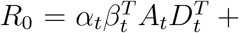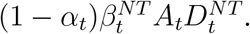

#### Parameter estimation

We infer model epidemiological parameters using an iterated filtering (IF) approach ((*1*)). In our model, we consider the unreported infection may be tested and reported later, so we add 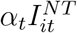 source code. We divided the full sample from January 1 and March 15 into five subperiods: January 10 to January 23, January 24 to February 2, February 3 to February 12, February 13 to February 22, and February 23 to March 15. We estimated the key parameters (*α, βdocumented, βundocumented, γdocumented, γundocumented*) for each period. The first period spans from the first day of Spring Festival to the day of Wuhan lockdown; the following three periods cover each of a 10-day interval up to February 22. After February 22, daily new cases dropped to a new level, and most of the containment policies were relaxing, so we set February 22 to March 15 as the final period.

Similar to (*1*)’s algorithm, the core model structure (Equations 1–5) was integrated stochastically using a 4th order Runge-Kutta (RK4) scheme. Specifically, for each step of the RK4 scheme, each unique term on the right-hand side of Equations 1–4 was determined using a random sample from a Poisson distribution. The initial values of the parameters were drawn using Latin hypercube sampling from uniform distributions with pre-specified ranges. The initial range for 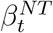 was set as 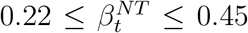 which is based on transmission rate range (i.e.[0.8,1.5] /ref) and mean value of within city flow 3.5. 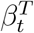 is equal to the transmission possibility of undetected infected patients multiply by a multiplicative factor 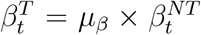. The initial ranges for *α_t_, µ_β_* and 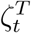 were chosen to cover most possible values, i.e. [0,1]. The initial ranges for the latency *Z_t_* were set from /ref 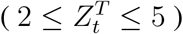. The initial ranges for the infection periods D in /ref is set as [2,5], since detection and treatment of infected patients may be reduce their infectious period, so we extend the range of the average duration of infection D to 1–5 days, where The average duration of infection for tested infected patients 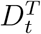 is 1–3 days and the average duration of infection for untested infected patients 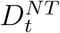 equal to 3–5 days.

#### Reporting delay

Cities in Hubei started to count clinically confirmed cases as confirmed COVID infections from February, 12. Patients who met clinical criteria through chest imaging but may not have had epidemiological links or a positive PCR test results were included in the official confirmed cases.^1^. As a direct consequence, the number of confirmed cases in Wuhan was more than 12 times higher than that in the previous day, creating a time-varying discrepancy in the case detection rate in Wuhan from other cities. To deal with it, we create a multiplication factor *µ_Wuhan, t_* for Wuhan and the detection rate in Wuhan was defined to be *µ_Wuhan,t_ ∗ α_t_*. To simulate the sharp increase case identification in Wuhan, we assume the reported rate of Wuhan equal to 1 on February 12. The initial exposed population*E_wuhan_* and initial undocumented infected population, *I_wuhan_* are were from a uniform distribution [0,*Seed_max_*]. *Seed_max_* was estimated at [1000,4000] in January 10 ((*1*)), and we compared the fitting results under different initial values, and found that *Seed_max_* = 3000 is the best fitting value (S6). We set the initial exposed population and initial undocumented infected population of other cities based on the number of travellers from Wuhan to city i on the first day of Chunyun. 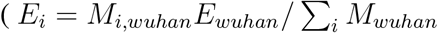 and 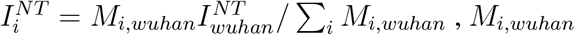 denotes the number of travelers from Wuhan to city i) In our model, we also considered a reported delay for tested infection. In China, cases are classified as suspected before reported officially as confirmed case, before that they must be tested at least two times. Suspected cases are sent to designated hospitals and quarantined before official confirmation^2^. Therefore, reported delay refers to the time interval between a person was admitted by a hospital and the observed confirmation of that individual infected case. In reality, many cases were confirmed after multiple tests, and the supply of testing reagent was insufficient at the beginning of the pandemic for timely confirmation.

We calculated the reported delay in our paper in a slightly different way from the approach employed in (*1*)^3^. To estimate this delay period, we relied on individual-level panel data compiled by a real-time epidemiological dataset^4^. We identified a trend break in report dealy after February 1 (S7). Therefore, we simulated the confirmed delay before and after February 1 We found that the time interval could be fit by the Gamma distribution(a = 3.55, b = 1.22, LL = 1669.71) before 2.1 and a = 3.86, b = 0.78, LL = 522.82 after 2.1) (S8). Since Hubei province included all clinically diagnosed cases to confirmed infections, we set the reported delay in Hubei equal to 0 in 2.12.

### Data

#### Observations of Confirmed COVID-19 Cases

We have compiled a city-level health outcome dataset in China for 339 cities from January 10, 2020, to March 15, 2020. From January 24, 2020, onwards, data were obtained from the public dataset Ding Xiang Yuan (DXY) that reports daily statistics across Chinese cities^5^. We used a web scraper program to obtain data from DXY 2–4 times every day. Data before January 24 can be obtained from the official website or official Weibo of National and Provincial Health Commissions in China.

#### Intercity Mobility data

We obtain inter-city population migration data from Baidu Migration^6^, a travel map offered by the largest Chinese search engine, Baidu. They calculate population flow based on Baidu mapping app user’s location and show the trajectory and characteristics of the population migration on the platform. For each of the 365 Chinese cities, the Baidu migration data reports the population inflow from the top 100 origin cities and outflow to the top 100 destination cities between January 21 and March 23 in 2019, and between January 10 and March 15 in 2020.^7^. In our main analysis, we rely on inter-city migration in 2019 to simulate the counterfactual spatial transmission of COVID-19 without traffic bans. Naturally, the reduction in intercity mobility in 2020 from its 2019 level is a combination of policy effects and individuals’ voluntary avoidance behaviour as a result of increased awareness. Our analysis is going to capture the composite impact of these two channels.

#### Within-city mobility data

Apart from the intercity data, Baidu also provides the daily within-city mobility data for each city in the sample period from a separate data product. The data is generated based on Baidu Map app usage within a city. We rely on this data to describe within-city mobility. Since Baidu’s app may not cover all population, we compare it to the mobility data in (*13*), which used nationwide mobile phone data to track population outflow from Wuhan from January 1 to January 24. The population mobility measured by mobile phone data sources is 5.5–6.5 times of the Baidu index. Therefore, We multiply inter-city mobility measures from Baidu by the multiplication factor of 6.

**Figure S1:**
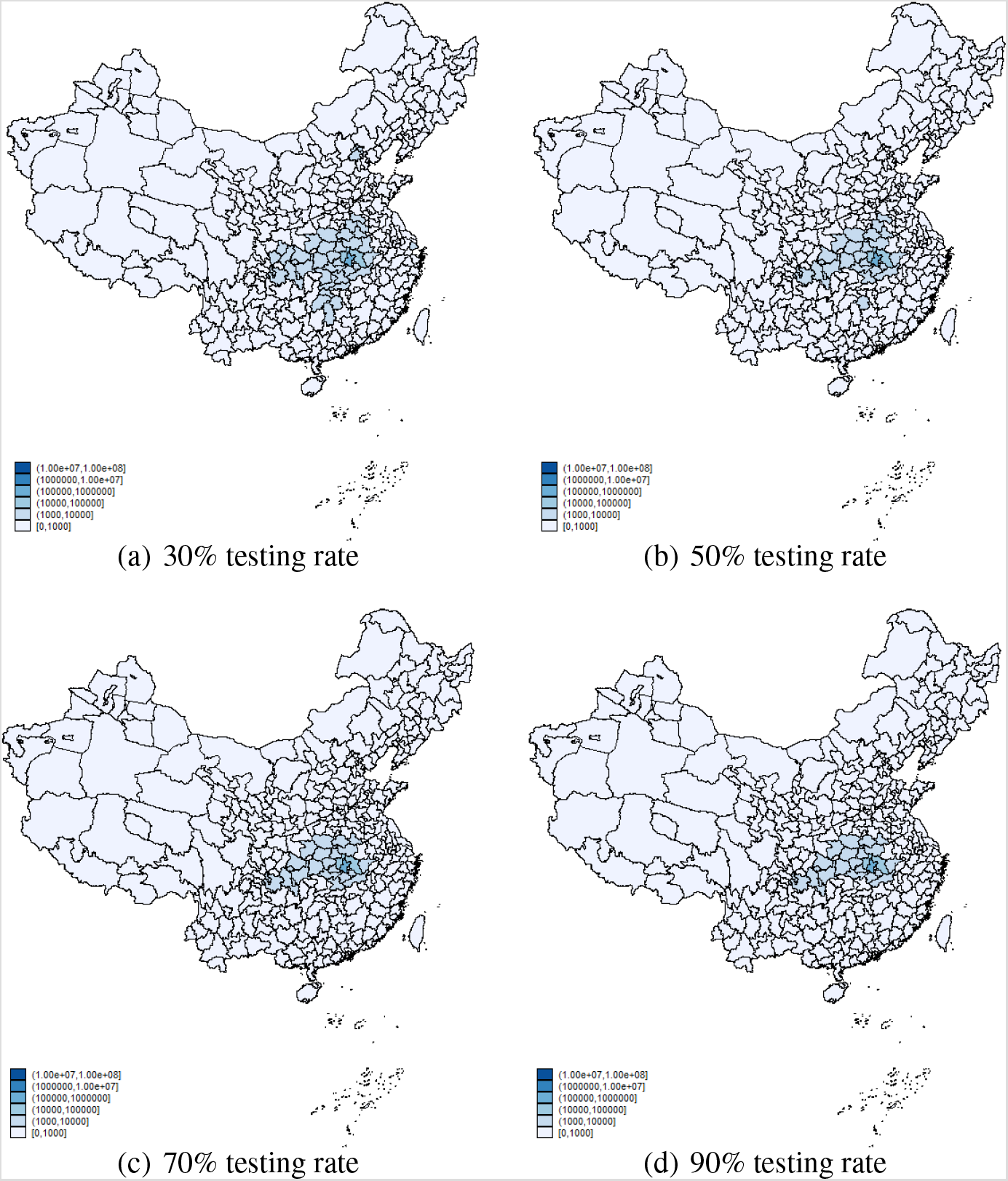
Spatial distribution of infection with different testing rate: keep lockdowns.

*Notes:* This graphs show spatial distribution of cumulative infections in March 15,2020 with both lockdown and intercity travel ban implemented (inter-city and within-city mobility were kept at 2020 level). Figure (a),(b),(c) and (d) shows the cumulative number of cases under different detection rate(30%,50%,70%, 90%).

**Figure S2:**
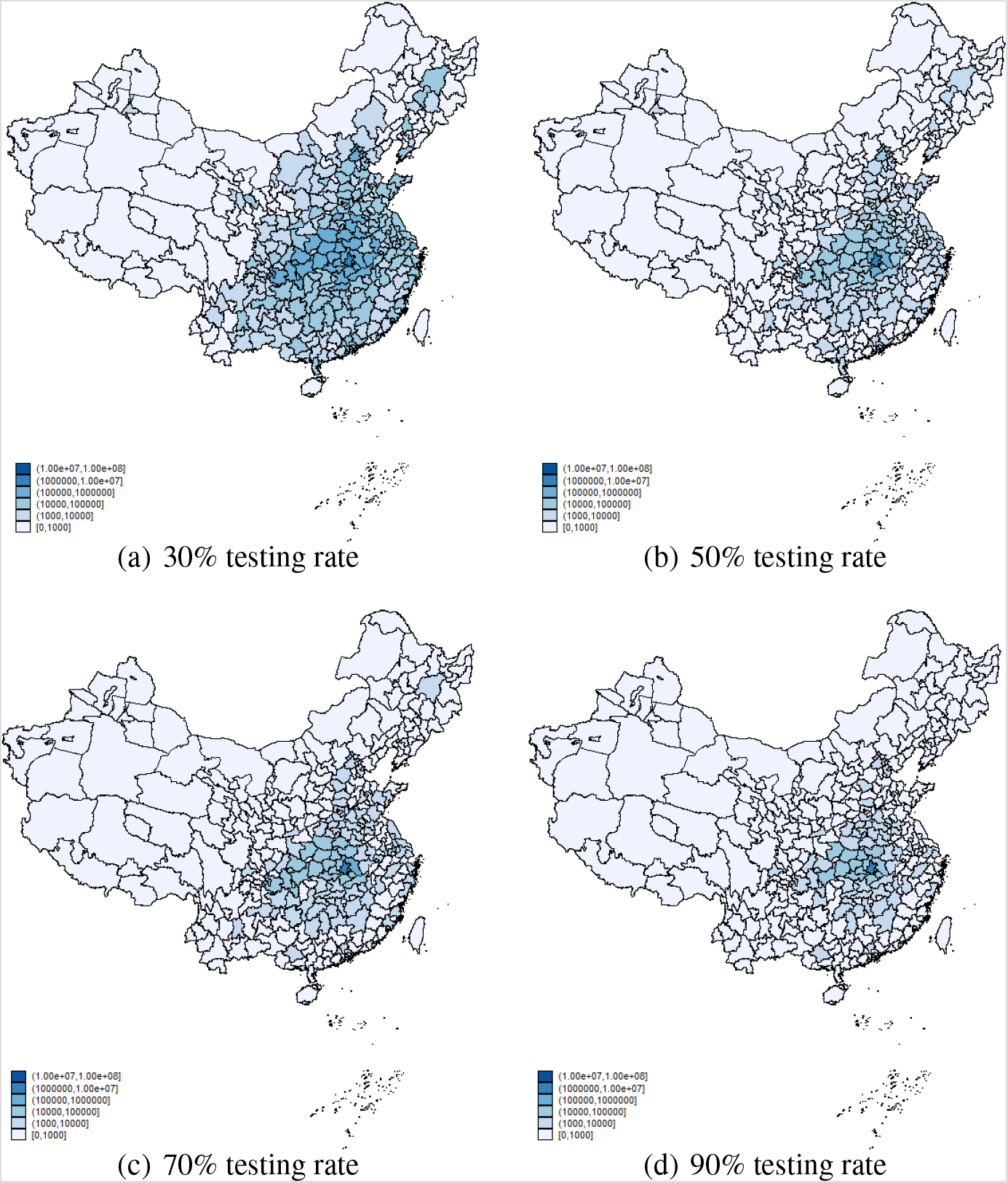
Spatial distribution of infection with Different Testing rate: drop lockdowns.

*Notes:* This graphs show spatial distribution of cumulative infections in March 15,2020 with both lockdown and intercity travel ban lifted (inter-city and within-city mobility were set at 2019 level). Figure (a),(b),(c) and (d) shows the cumulative number of cases under different detection rate(30%,50%,70%, 90%).

**Figure S3:**
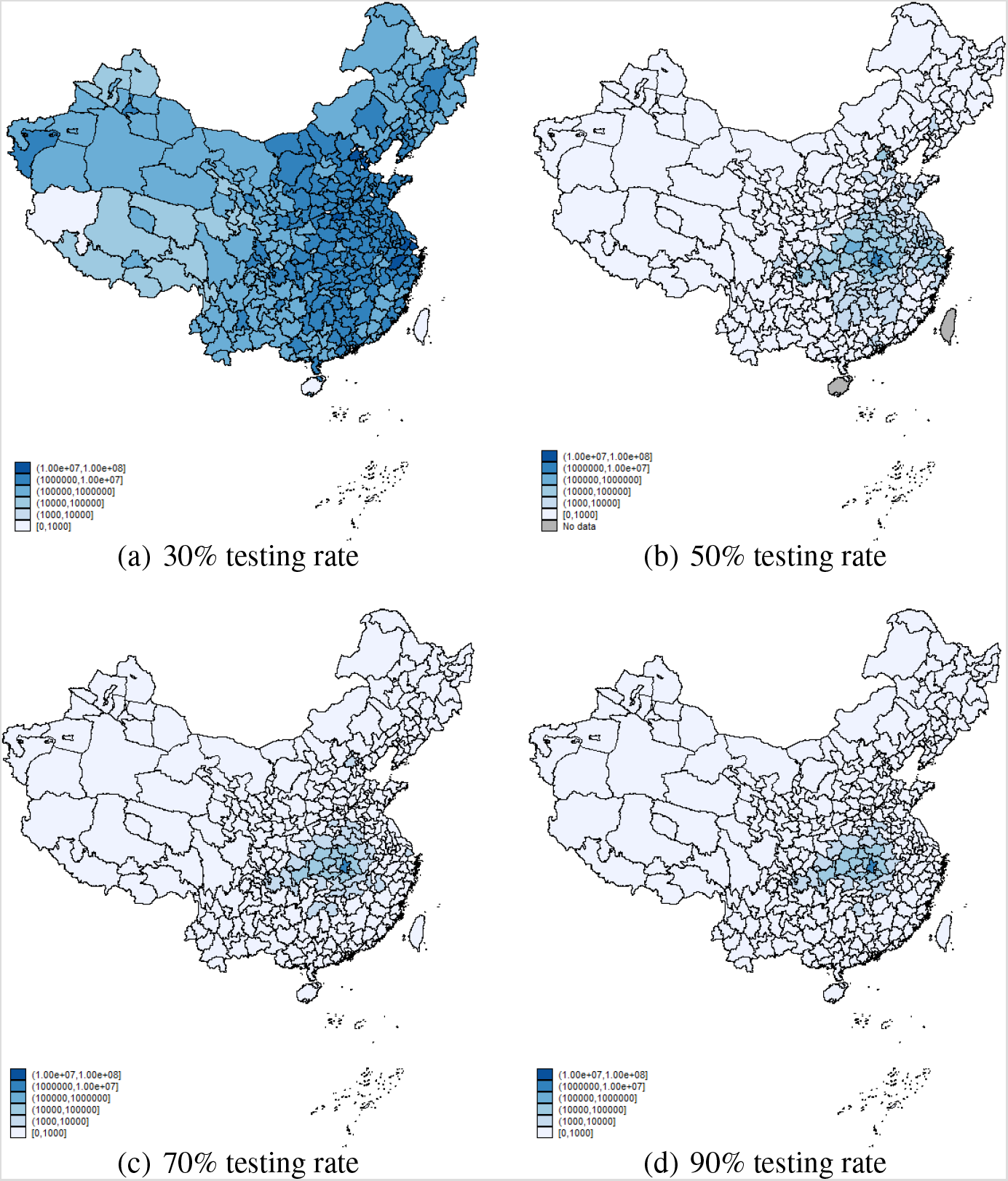
Prediction of Spatial distribution of infection with different Testing rate: keep lockdown.

*Notes:* This graphs show spatial distribution of cumulative infections in July 15,2020 with both lockdown and intercity travel ban implemented (inter-city and within-city mobility were set at 2020 level). Figure (a),(b),(c) and (d) shows the cumulative number of cases under different detection rate (30%, 50%, 70%, 90%).

**Figure S4:**
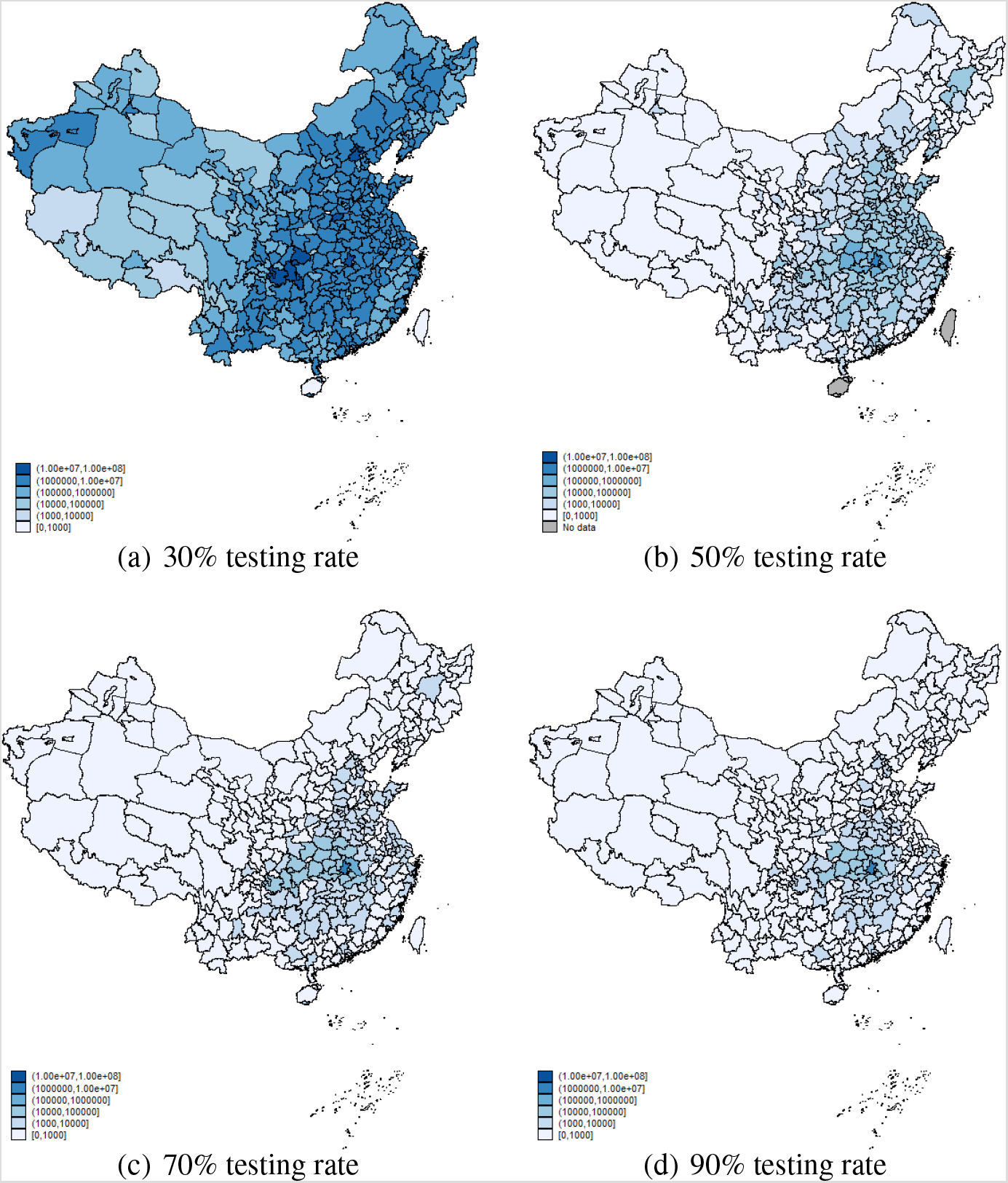
Prediction of Spatial distribution of infection with Different Testing rate: drop lockdown.

*Notes:* This graphs show spatial distribution of cumulative infections in J 15,2020 with both lockdown and intercity travel ban lifted (inter-city and within-city mobility were set at 2019 level). Figure (a),(b),(c) and (d) shows the cumulative number of cases under different detection rate(30%,50%,70%, 90%).

**Figure S5:**
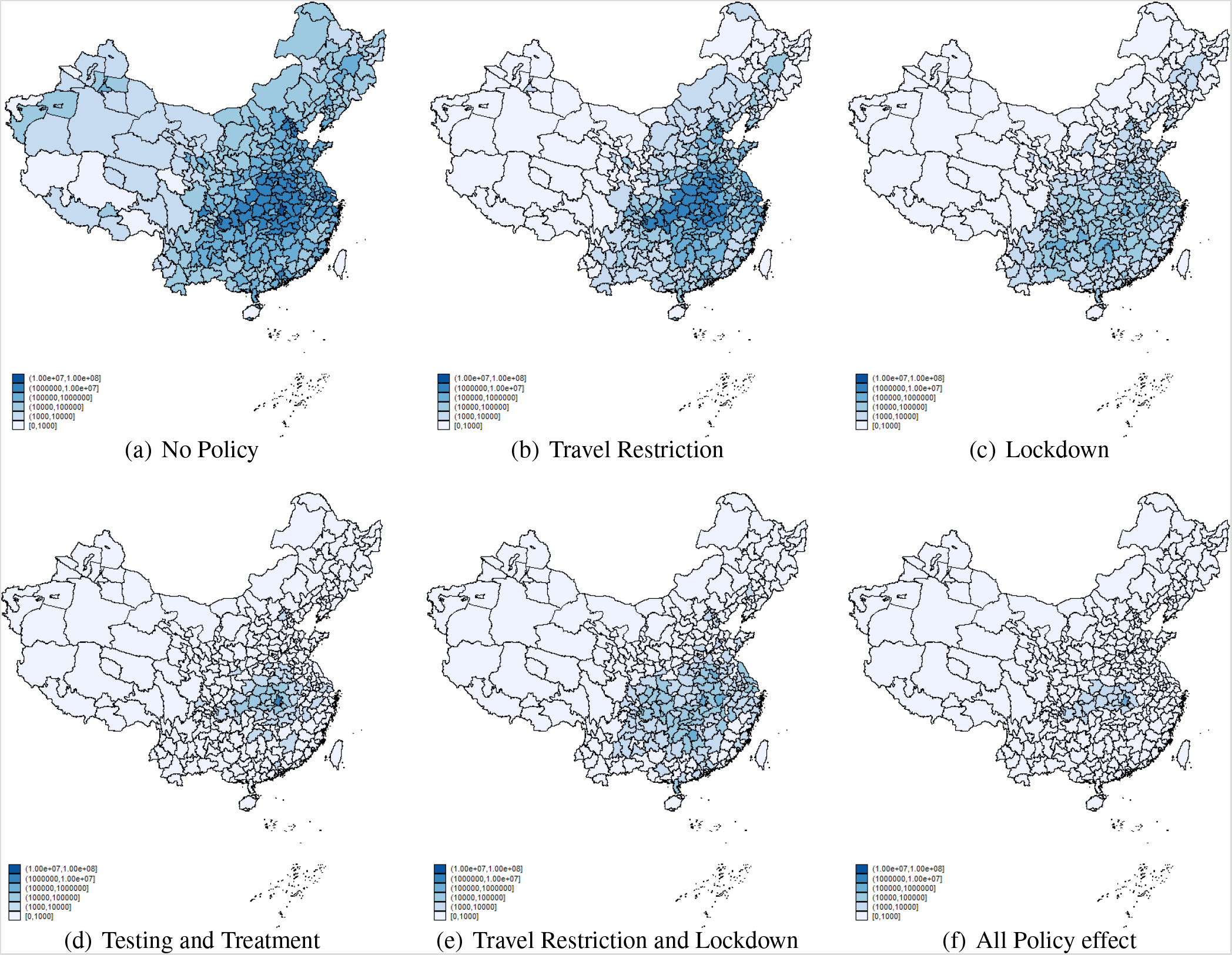
Spatial distribution of infections under alternative scenarios: March 15, 2020.

*Notes:* This graph shows the spatial distribution of cumulative infectious cases on March 15, 2020 under different scenarios.Figure (a) plots number of total reported case distribution without any policy implemented. Figure (b),(c),(d) and (e) shows the cumulative number of cases under travel restriction, lock-down, testing and treatment and the combination of travel restriction and lock-down,respectively. Figure (f) simulates the baseline distribution of infections with all three policies in place.

**Figure S6:**
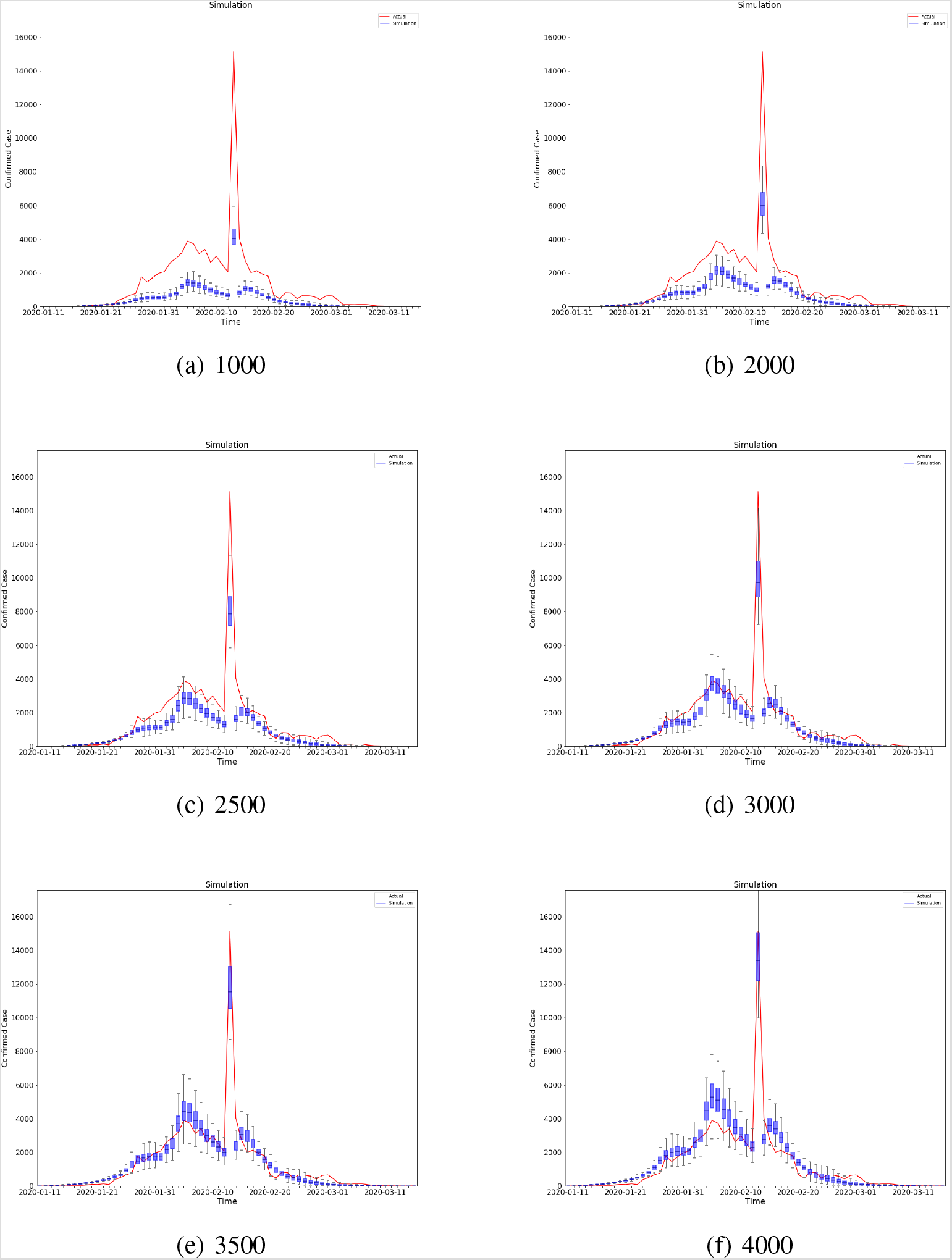
Simulations with different initial values.

Notes: The graphs plot simulations using different initial value. The observed cases is best fitted when initial value equal to 3000 (MAE = 363) than other initial values (*MAE*_*seed*=1000_ = 848,*MAE*_*seed*=1500_ = 660, *MAE*_*seed*=2000_ = 489, *MAE*_*seed*=2500_ = 373, *MAE*_*seed*=3000_ = 354,*MAE*_*seed*=3500_ = 438, *MAE*_*seed*=4000_ = 554.

**Figure S7:**
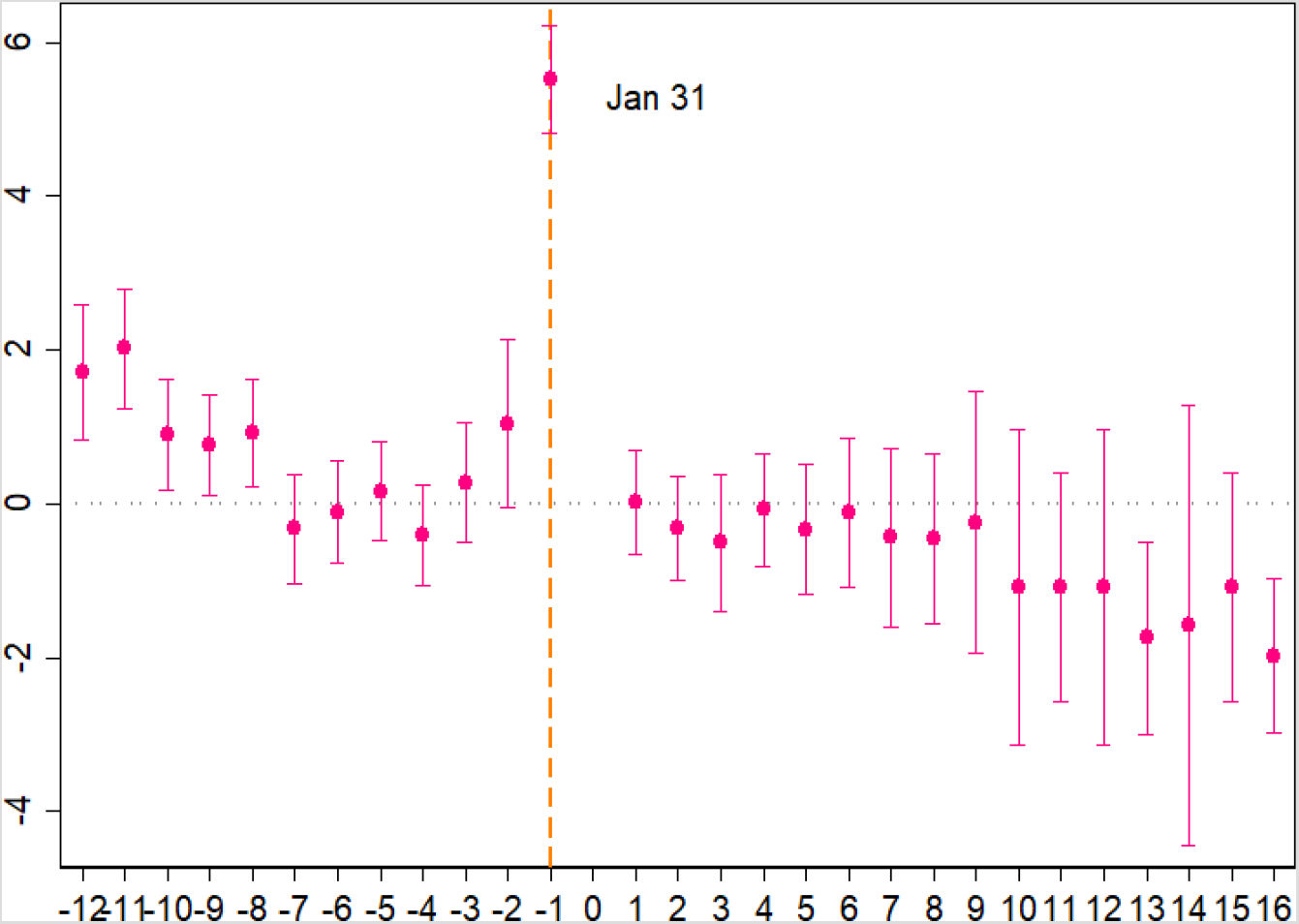
Reporting delay.

*Notes:* t0 is set to be January 31, 2020. We observed the change of reporting delay between January 18 and February 15 days in which hospital registration data were available.

**Figure S8:**
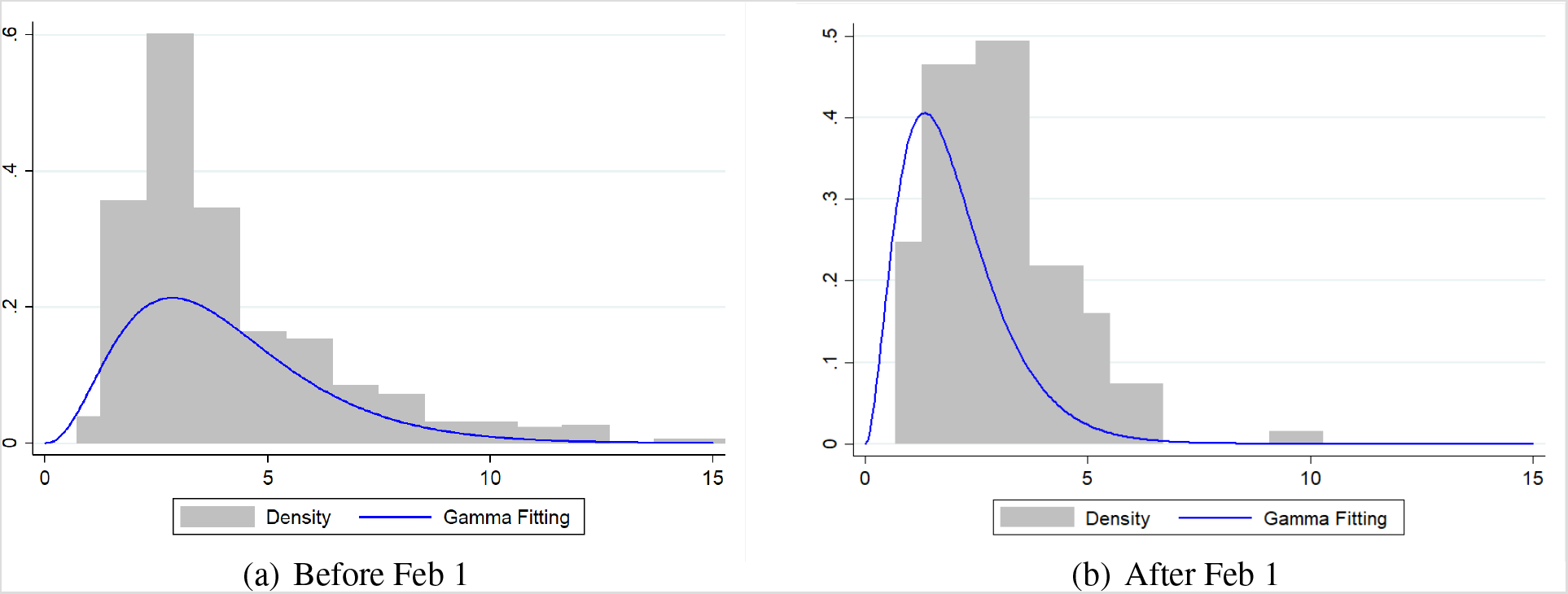
Reporting delay: Gamma function.

*Notes:* The graphs plot the histograms of case reporting delay for cases confirmed before and after February 1. Reporting delay is defined as the interval between the day each admitted by the hospital and the day each case was confirmed officially. The data prior to February 1 were fitted with a Gamma distribution(a = 3.55, b = 1.22, LL = 1669.71) and data were fitted with a Gamma distribution (a = 3.86, b = 0.78, LL = 522.82 after 2.1) after February 1,2020.

## Footnotes

1 http://wjw.hubei.gov.cn/fbjd/dtyw/202002/t20200213_2025581.shtml. InFebruary20,Hubeicancelledtheclinicalconfirmmethod:http://news.jcrb.com/jxsw/202002/t20200220_2116026.html

2 Details are in Diagnosis and Treatment Protocol for COVID-19, which published by National Health Commission of the People’s Republic of China. Source: http://www.gov.cn/zhengce/zhengceku/2020-02/05/5474791/files/de44557832ad4be1929091dcbcfca891.pdf

3 They accounted for the time interval between a person transitioning from latent to contagious and the observational confirmation of that individual infection.

4 Data Source: https://github.com/beoutbreakprepared/nCoV2019

5 https://ncov.dxy.cn/ncovh5/view/pneumonia

6 Source: http://qianxi.baidu.com/

7 Baidu migration mostly serves to show population flow during the Spring Festival, so the available period of data is mainly based on the time of Chinese New Year according to the lunar calendar.

## Notes

### Competing Interest Statement

The authors have declared no competing interest.

### Author Declarations

no IRB needed

